# Vitamin B12 in pregnancy and its relationship with maternal BMI and gestational DM

**DOI:** 10.1101/2023.02.16.23286019

**Authors:** Rameesha Muzaffar, Jameeha Khursheed, Anum Yousaf

## Abstract

**Introduction:** Gestational diabetes mellitus (GDM) is one of the most prevalent complications of pregnancy. In a joint statement released in 2018, the International Federation of Obstetrics and Gynecology (FIGO) and the International Diabetes Federation (IDF) stated that high blood glucose levels during pregnancy have an impact on maternal, newborn, and child health and may contribute to the global burden of type 2 diabetes mellitus and cardiovascular metabolic disorders in the short and long term.

**Objectives:** The basic aim of the study is to analyze vitamin B12 in pregnancy and its relationship with maternal BMI and gestational DM.

**Material and methods:** This cross-sectional study was conducted in THQ Hospital Deepalpur, Pakistan during March 2021 to August 2021. Record gathered from 118 hospitals with the total number of 364 women stated for the study coordinator. As per database record, 11% (41 women) were excluded due to initial trimester spontaneous abortion as well as type 2 diabetes diagnosed in 16; 4% and follow up loss 2; 1%.

**Results:** Maternal outcomes in pregnant women with type 1 diabetes and those without the disease were evaluated in this study. No maternal mortality occurred within 30 days of delivery in 630 pregnancies with type 1 diabetes. However, pregnant women with type 1 diabetes were usually at a much higher risk of developing adverse maternal events during their pregnancy than women without type 1 diabetes, even after adjusting for age and infant sex or age, infant sex, place of residence, income level, occupation, calendar year, and Charlson comorbidity index.

**Conclusion:** It is concluded type 1 diabetes remains a significant disease threatening pregnant women and their offspring. Clinicians should be aware of this clinical situation.

## Introduction

Gestational diabetes mellitus (GDM) is one of the most prevalent complications of pregnancy. In a joint statement released in 2018, the International Federation of Obstetrics and Gynecology (FIGO) and the International Diabetes Federation (IDF) stated that high blood glucose levels during pregnancy have an impact on maternal, newborn, and child health and may contribute to the global burden of type 2 diabetes mellitus and cardiovascular metabolic disorders in the short and long term. This poses a critical public health challenge as well as a substantial financial burden. Type 1 diabetes holding pregnant women are also associated with a highly increased risk of congenital malformations, neonatal morbidity, and obstetric complications. These highly adverse results are related to pre-conceptional care typically related to the glycaemic control level. Sufficient pre-conceptional care declines the congenital malformation frequency and boosts the pregnancy result [1].

Encouraging diabetic women to manage their pregnancies; to begin supplements of folic acid; to optimize in the control of glycaemic before conception; is however considered as a recognized objective [2]. Our gathered data is based on the PubMed Database, collected through different centers with specific thought of pregnancy and diabetes, but not associated with the total local population. Nationwide population data are infrequent and most of the data have been gathered retrospectively [3].

The pregnancy outcomes in type 1 diabetic patients reported between 1991 and 2003 are grim. High rates of maternal and fetal adverse outcomes highlight the challenges physicians face in the management of pregnant patients with the disease [4]. The causes of adverse pregnancy outcomes may include a high proportion of unplanned pregnancies with prior poor glycemic control, failure to maintain tight metabolic control during pregnancy, and pre-existing medical comorbidities, such as nephropathy and hypertension [5].

Care for type 1 diabetes has advanced with time. Preconception and early pregnancy care to achieve optimal glycemic control has lowered perinatal mortality and the rate of fetal malformations. The widespread adoption of blood glucose self-monitoring has enabled patients to adjust their insulin dose in a timely manner and modify their lifestyle to control their glucose levels [6]. Compared with human insulin, a fast-acting insulin analogue has been demonstrated to lower the risk of fetal adverse outcomes. New insulin delivery systems, including continuous subcutaneous insulin infusion, sensor-augmented pump therapy, and closed-loop insulin delivery, have also provided additional tools to improve glycemic control. In addition to glycemic control, multidisciplinary patient-centered care of type 1 diabetes before and during pregnancy likely has led to better gestational outcomes [7].

### Objectives

The basic aim of the study is to analyze vitamin B12 in pregnancy and its relationship with maternal BMI and gestational DM.

## Material and methods

This cross-sectional study was conducted in THQ Hospital Deepalpur, Pakistan during March 2021 to August 2021. Record gathered from 118 hospitals with the total number of 364 women stated for the study coordinator. As per database record, 11% (41 women) were excluded due to initial trimester spontaneous abortion as well as type 2 diabetes diagnosed in 16; 4% and follow up loss 2; 1%. We also stated the assessment of 323 pregnancies and all respondents gave written apprised consent. Entitled women completed questionnaires at inclusion (at the end of the initial trimester and around gestation of ten weeks) but during the trimester three (which is almost around 34 weeks). Internists complete the specific abovementioned questionnaire comprising common features, history of medical and other those items which are diabetes related; accordingly, gynecologists provided information regarding the pregnancy results and finally pediatricians completed in a specific questionnaire to gather newborns’ information. We also gather (through PubMed Database) BMI, age, marital status, level of education, ethnic origin, use of alcohol, parity and smoking habits.

### Statistical analysis

The data were sampled and entered into the SPSS worksheet for analysis. The alpha criterion was set at 0.05. After constructing a 2×2 contingency table, chi-square without Yates correction was used to find the association between the potential risk factors and pregnancy status.

## Results

Maternal outcomes in pregnant women with type 1 diabetes and those without the disease were evaluated in this study. No maternal mortality occurred within 30 days of delivery in 630 pregnancies with type 1 diabetes. However, pregnant women with type 1 diabetes were usually at a much higher risk of developing adverse maternal events during their pregnancy than women without type 1 diabetes, even after adjusting for age and infant sex or age, infant sex, place of residence, income level, occupation, calendar year, and Charlson comorbidity index. The risks of preeclampsia, eclampsia and cesarean delivery, increased in the type 1 diabetes cohort. Pregnant women with type 1 diabetes were at a higher risk of developing pregnancy-related hypertension, puerperal cerebrovascular disorders, acute renal failure, shock, intracranial injuries, cardiac arrest/ventricular fibrillation, acute myocardial infarction, severe anesthesia complications, and thrombotic embolism.

**Table 01:**
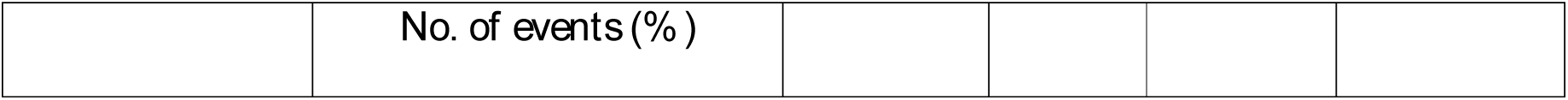
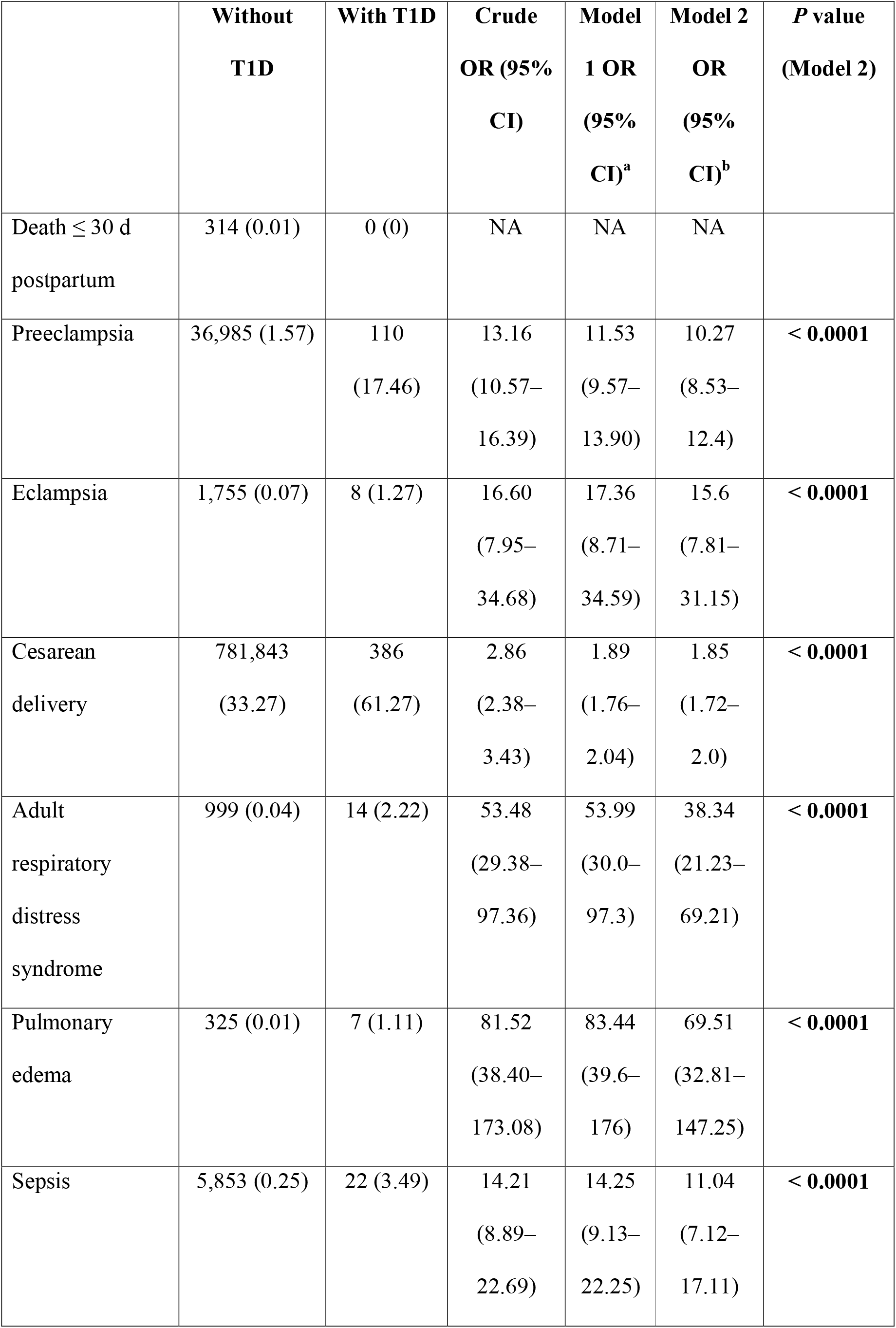
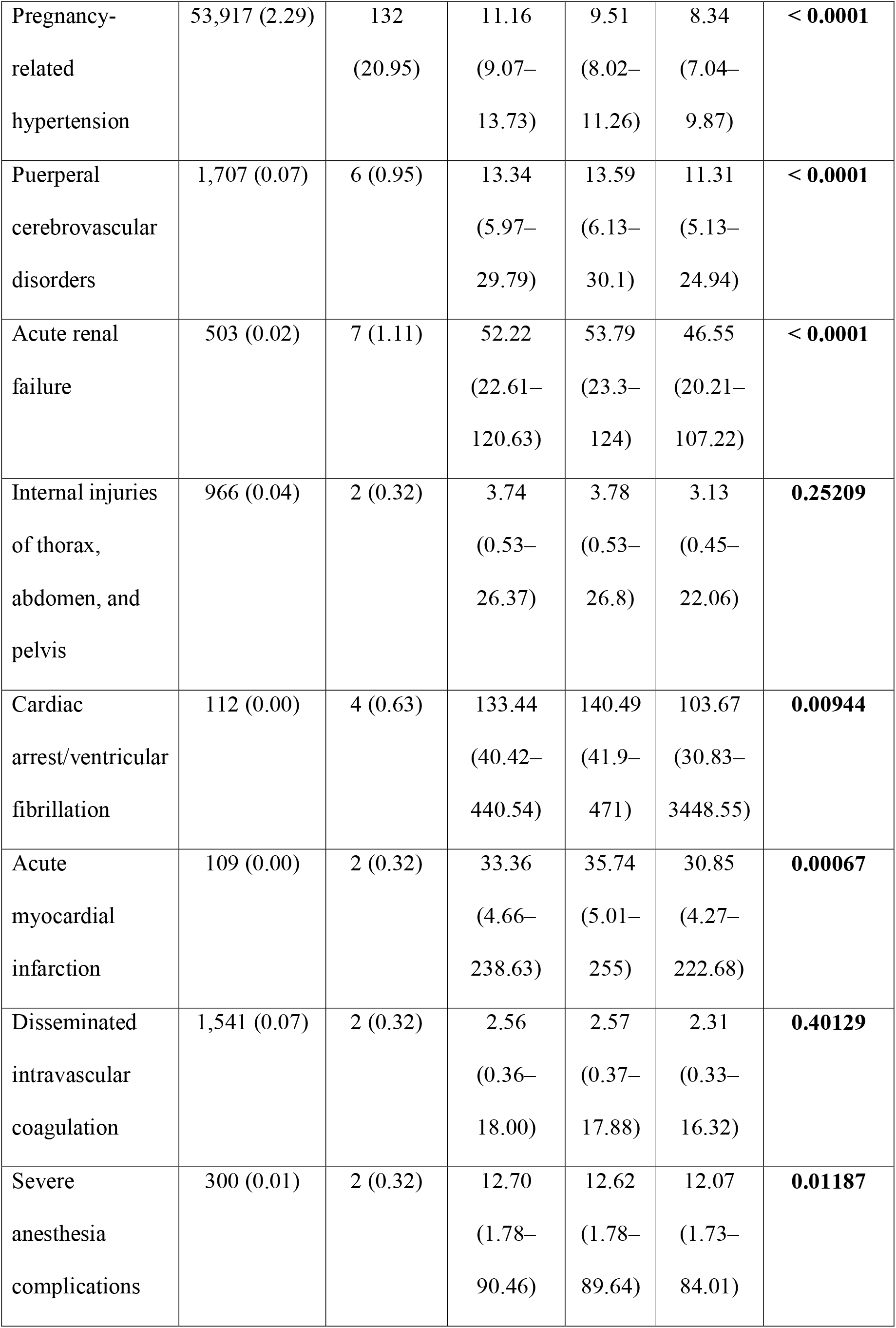
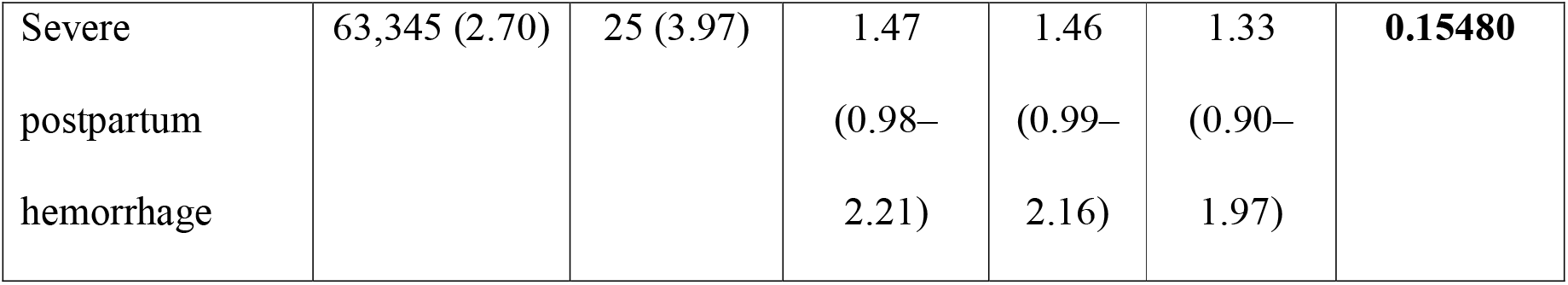
Maternal outcomes among pregnant women without type 1 diabetes and pregnant women with type 1 diabetes

|  | **Without**<br>**T1D** | **With T1D** | **Crude**<br>**OR (95%**<br>**CI)** | **Model**<br>**1 OR**<br>**(95%**<br>**CI)<sup>a</sup>** | **Model 2**<br>**OR**<br>**(95%**<br>**CI)<sup>b</sup>** | **P value**<br>**(Model 2)** |
| --- | --- | --- | --- | --- | --- | --- |
| Death ≤ 30 d<br>postpartum | 314 (0.01) | 0 (0) | NA | NA | NA |  |
| Preeclampsia | 36,985 (1.57) | 110<br>(17.46) | 13.16<br>(10.57–<br>16.39) | 11.53<br>(9.57–<br>13.90) | 10.27<br>(8.53–<br>12.4) | **< 0.0001** |
| Eclampsia | 1,755 (0.07) | 8 (1.27) | 16.60<br>(7.95–<br>34.68) | 17.36<br>(8.71–<br>34.59) | 15.6<br>(7.81–<br>31.15) | **< 0.0001** |
| Cesarean<br>delivery | 781,843<br>(33.27) | 386<br>(61.27) | 2.86<br>(2.38–<br>3.43) | 1.89<br>(1.76–<br>2.04) | 1.85<br>(1.72–<br>2.0) | **< 0.0001** |
| Adult<br>respiratory<br>distress<br>syndrome | 999 (0.04) | 14 (2.22) | 53.48<br>(29.38–<br>97.36) | 53.99<br>(30.0–<br>97.3) | 38.34<br>(21.23–<br>69.21) | **< 0.0001** |
| Pulmonary<br>edema | 325 (0.01) | 7 (1.11) | 81.52<br>(38.40–<br>173.08) | 83.44<br>(39.6–<br>176) | 69.51<br>(32.81–<br>147.25) | **< 0.0001** |
| Sepsis | 5,853 (0.25) | 22 (3.49) | 14.21<br>(8.89–<br>22.69) | 14.25<br>(9.13–<br>22.25) | 11.04<br>(7.12–<br>17.11) | **< 0.0001** |
| Pregnancy-related hypertension | 53,917 (2.29) | 132<br>(20.95) | 11.16<br>(9.07–13.73) | 9.51<br>(8.02–11.26) | 8.34<br>(7.04–9.87) | **< 0.0001** |
| Puerperal cerebrovascular disorders | 1,707 (0.07) | 6 (0.95) | 13.34<br>(5.97–29.79) | 13.59<br>(6.13–30.1) | 11.31<br>(5.13–24.94) | **< 0.0001** |
| Acute renal failure | 503 (0.02) | 7 (1.11) | 52.22<br>(22.61–120.63) | 53.79<br>(23.3–124) | 46.55<br>(20.21–107.22) | **< 0.0001** |
| Internal injuries of thorax, abdomen, and pelvis | 966 (0.04) | 2 (0.32) | 3.74<br>(0.53–26.37) | 3.78<br>(0.53–26.8) | 3.13<br>(0.45–22.06) | **0.25209** |
| Cardiac arrest/ventricular fibrillation | 112 (0.00) | 4 (0.63) | 133.44<br>(40.42–440.54) | 140.49<br>(41.9–471) | 103.67<br>(30.83–3448.55) | **0.00944** |
| Acute myocardial infarction | 109 (0.00) | 2 (0.32) | 33.36<br>(4.66–238.63) | 35.74<br>(5.01–255) | 30.85<br>(4.27–222.68) | **0.00067** |
| Disseminated intravascular coagulation | 1,541 (0.07) | 2 (0.32) | 2.56<br>(0.36–18.00) | 2.57<br>(0.37–17.88) | 2.31<br>(0.33–16.32) | **0.40129** |
| Severe anesthesia complications | 300 (0.01) | 2 (0.32) | 12.70<br>(1.78–90.46) | 12.62<br>(1.78–89.64) | 12.07<br>(1.73–84.01) | **0.01187** |
| Severe<br>postpartum<br>hemorrhage | 63,345 (2.70) | 25 (3.97) | 1.47<br>(0.98–<br>2.21) | 1.46<br>(0.99–<br>2.16) | 1.33<br>(0.90–<br>1.97) | **0.15480** |

## Discussion

This study showed strong correlations between type 1 diabetes and multiple adverse outcomes, including pregnancy-related hypertension, preeclampsia, eclampsia, cesarean delivery, stillbirth, and preterm birth. In addition to these well-recognized adverse outcomes, we also found that patients with type 1 diabetes were associated with high risks of other major morbidities, including adult respiratory distress syndrome, pulmonary edema, sepsis, chorioamnionitis, puerperal cerebrovascular disorders, acute renal failure, and shock [8]. These data emphasize that type 1 diabetes is associated with increased adverse outcomes in both mothers and fetuses.

We found that the only improvement in outcome was a lower risk of preeclampsia in the late period as compared with that in the early period in pregnant women with type 1 diabetes. The reasons behind for this improvement are unclear. Better glucose and blood pressure control, dietary and lifestyle modification, low-dose aspirin, and calcium supplementation in women with low dietary calcium intake have been reported to reduce the risk of preeclampsia [9]. We further supposed that the availability of free glucose test strips had relieved the financial stress associated with the disease and led to better diabetes care and outcomes. The decreased risk of preeclampsia is of high clinical significance because preeclampsia is associated with maternal morbidity and mortality during pregnancy and is a strong predictor of future cardiovascular disease in mothers [10].

## Conclusion

It is concluded type 1 diabetes remains a significant disease threatening pregnant women and their offspring. Clinicians should be aware of this clinical situation.

## Data Availability

All data produced in the present work are contained in the manuscript

